# Evaluating the Efficacy of Large Language Models in Addressing Patient-Centric Inquiries in Multiple Cancers

**DOI:** 10.1101/2025.08.05.25332968

**Authors:** Soheila Borhani, Xiaoqian Jiang

## Abstract

**Background:** Large Language Models (LLMs) have transformed how patients access health information online. Chatbots like ChatGPT allow users to ask direct questions and receive tailored answers almost instantly. However, for LLMs to be effective, the answers they provide must be reliable and accessible to patients. Our review assessed the reliability and accessibility of LLMs in answering patient inquiries about breast, prostate, and lung cancer.

**Methods:** A systematic search of the PubMed, Embase, and Web of Science databases was conducted. Included studies were peer-reviewed original research, published in English, that evaluated one or more LLMs in answering patients’ oncology questions. To enable result aggregation, a linear transformation was applied to standardize data from studies that used different Likert scales.

**Results:** We identified three common measures of reliability (accuracy, quality, consistency), and three measures of accessibility (readability, understandability, actionability) across the thirty-six studies that met our inclusion criteria. Accuracy and quality scores showed roughly similar distributions, with median values of 79.0% and 76.5%, respectively. Consistency levels were high in the few studies that provided this data (median = 100%). Despite all LLMs having readability scores significantly below the recommended level for patient-facing materials (median = 40.4%), several studies reported substantial improvements through prompt engineering. Understandability (median = 69.0%) and particularly, actionability (median = 40.0%) scores were lower than desired.

**Conclusions:** Despite current limitations, LLMs hold significant potential as an assistant tool for disseminating health information to patients. Active involvement of physicians in model training and validation can help improve their performance.

## 1. Introduction

Patients are increasingly turning to the internet for health information, with Google alone processing over a billion health-related searches daily [1]. Queries related to cancer make up a notable portion of these searches, due to the high prevalence of cancer, and the anxiety that it evokes in patients. Nearly nine in ten (89%) cancer survivors reported searching for cancer information online after receiving their diagnosis [2]. However, the quality of information available online can be quite poor. A review of the most popular cancer-related articles on Facebook, Reddit, Twitter, and Pinterest found that over 30% of them contained misleading or harmful information [3]. A similar study [4] revealed that 90% of prostate cancer content on Instagram was of low to medium quality, with 40% containing significant misinformation.

Recent advances in Artificial Intelligence (AI), particularly in the domain of Large Language models (LLMs), present a paradigm shift in how patients access health information online. Rather than using search engines to navigate numerous links and synthesize information themselves, patients can pose direct questions to LLM-powered chatbots, and receive specific, detailed, and tailored answers in a matter of seconds. The advantage of LLMs over traditional search engines is underscored by the immense popularity and rapid adoption of this new technology. ChatGPT, the first widely accessible LLM-based chatbot developed by OpenAI Inc., achieved a record-breaking 100 million users within just two months of its launch in late 2022 [5]. Shortly after, other technology companies followed suit by releasing their own LLM-based chatbots, as summarized visually in the timeline shown in Figure 1.

**Figure 1.**
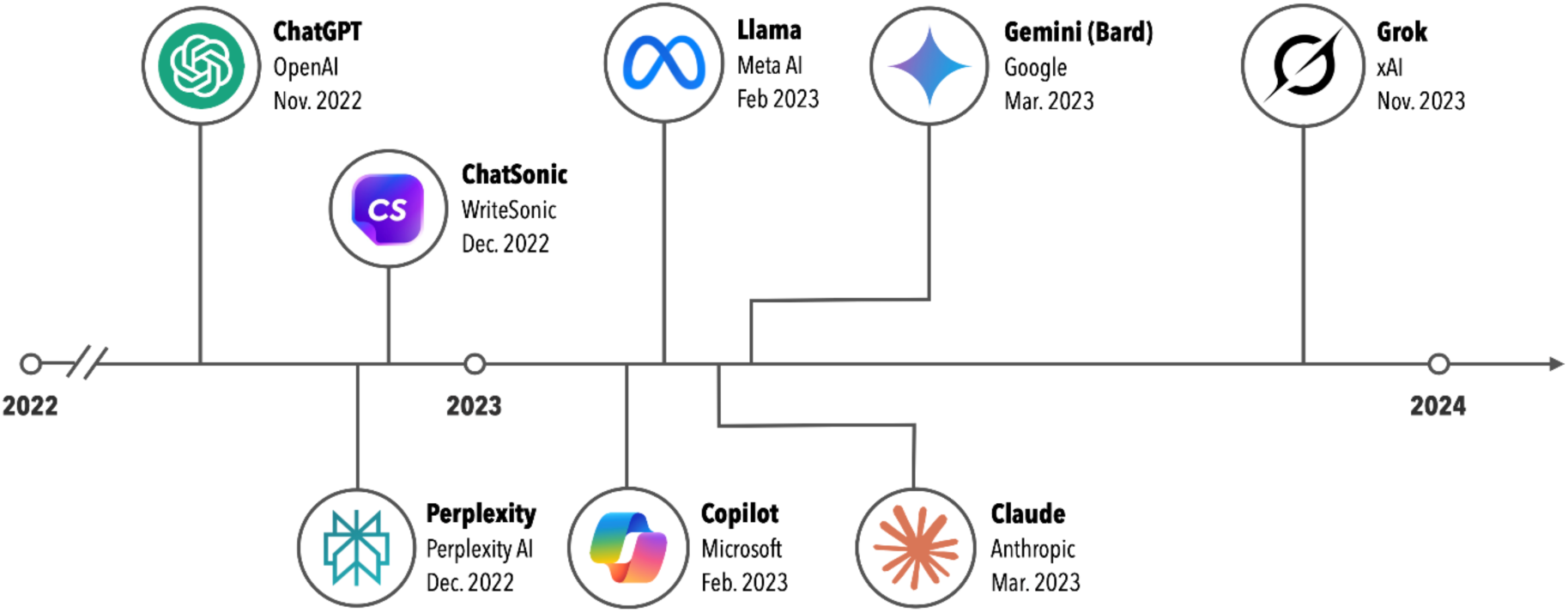
A timeline of the public releases of popular LLM-based chatbots.

Despite their transformative potential, LLMs remain susceptible to inaccuracies and misinformation [6,7]. As generative AI models, LLMs utilize deep neural network architectures with billions of tunable parameters. These parameters, algorithmically adjusted in a process called training, enable LLMs to identify statistical patterns and word relationships in written language [8]. The data used to train LLMs is sourced from a wide and diverse array of digital content including books, scientific papers, news articles, blogs, and social media posts, to name a few. LLMs can therefore inherit any inaccuracies or biases present in their training data [9]. For example, it is estimated that the majority of the data for training LLMs originates in the U.S. [10]. As a result, cancer screening recommendations may be skewed towards U.S. standards and guidelines, potentially providing misleading information to patients outside the United States [11]. Moreover, much of the information found online about a specific cancer drug or treatment may originate from the manufacturing company, rather than authoritative medical sources, which creates a real risk of commercial bias [12].

Considering the risk of inaccuracies and misinformation in LLMs, their successful use as a source of information for cancer patients depends entirely on the reliability of their output; otherwise, they could cause widespread harm through large-scale dissemination of false information. Reliability, however, is only a necessary condition not a sufficient one. To be beneficial to patients, the information must be presented in a manner that is easy to access and comprehend. In other words, LLM-generated content should be both *reliable* and *accessible*. Therefore, in this study we aimed to review the literature to determine whether the answers provided by LLMs to patients’ cancer-related questions were reliable and accessible. We focused on the three most commonly diagnosed cancers (i.e., breast, prostate, and lung cancer) which combined, account for over 40% of new cancer cases in the United States [13]. To our knowledge, this is the first systematic review to evaluate the performance of LLMs as a source of information for cancer patients.

## 2. Methods

Using the search string displayed in Figure 2, we conducted a search of article titles and abstracts indexed in three international databases, namely, PubMed, Embase, and Web of Science. Due to LLMs’ recency, we limited the scope of our search to studies published within the past 5 years. Initially, 496 articles were retrieved across all three databases. This number was subsequently reduced to 283 after removing duplicate records. Next, the titles and abstracts of the remaining articles were screened to exclude those that appeared prima facie irrelevant to the topic at hand. Also excluded were articles in languages other than English, publications that were not peer-reviewed, editorials, correspondences, and commentaries. The full text of the remaining articles was read to ensure they specifically addressed and assessed the use of at least one LLM in the context of answering patient questions. In total, thirty-six studies met the criteria described above, and were included in the final review. The selection process is summarized in the Preferred Reporting Items for Systematic Reviews and Meta-Analyses (PRISMA) diagram shown in Figure 2.

**Figure 2.**
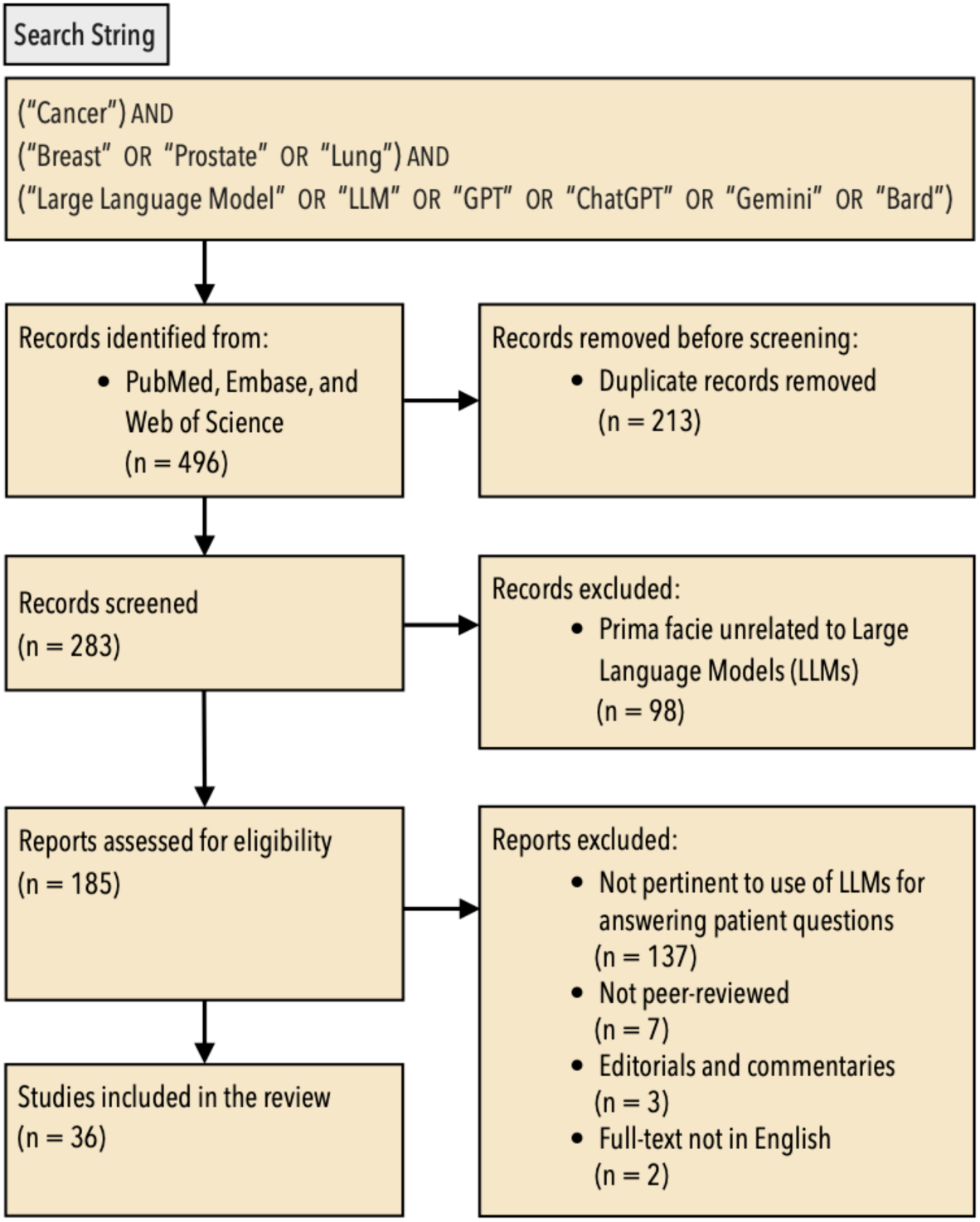
PRISMA flow chart summarizing the process of identification, screening, and selection of the studies included in the review.

Data extracted from the selected studies included: cancer site (e.g., breast, prostate, lung, or multiple), LLM type and version (e.g., ChatGPT-3.5), the questions posed to LLMs, the number and expertise of human evaluators, and how they graded the LLM responses. To enable result aggregation across studies employing different Likert scales, we applied a data conversion scheme by linearly mapping all scores onto a common 0-100 range, as described in the Results section in further detail. Python version 3.9.6 was used for linear regression analysis and data visualization.

## 3. Results

Table 1 presents an overview of the studies included in this review, detailing, among other information, the LLMs utilized in each study along with the number and source of the questions posed to them. These questions originated from a variety of sources, including the FAQ sections of the websites of various cancer associations (e.g., [40]), questions formulated by the authors based on their clinical oncology practice and experience (e.g., [46]), questions designed by focus groups of patient advocates (e.g., [35]), and questions identified by analyzing Google Trends data (e.g., [28]). Notably, in one study [19] the researchers tasked the LLM itself to generate the most common patient questions about prostate cancer.

**Table 1.**
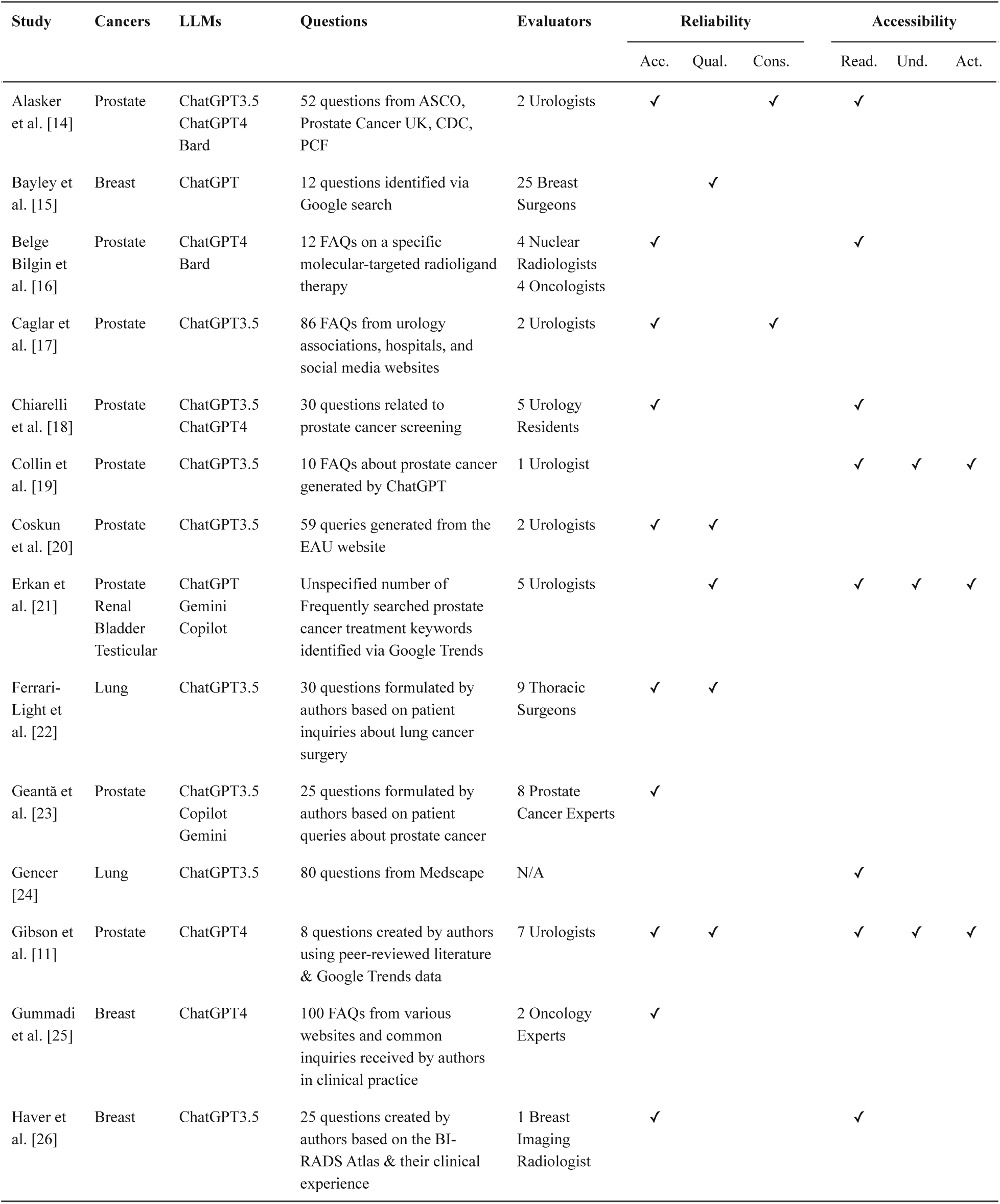
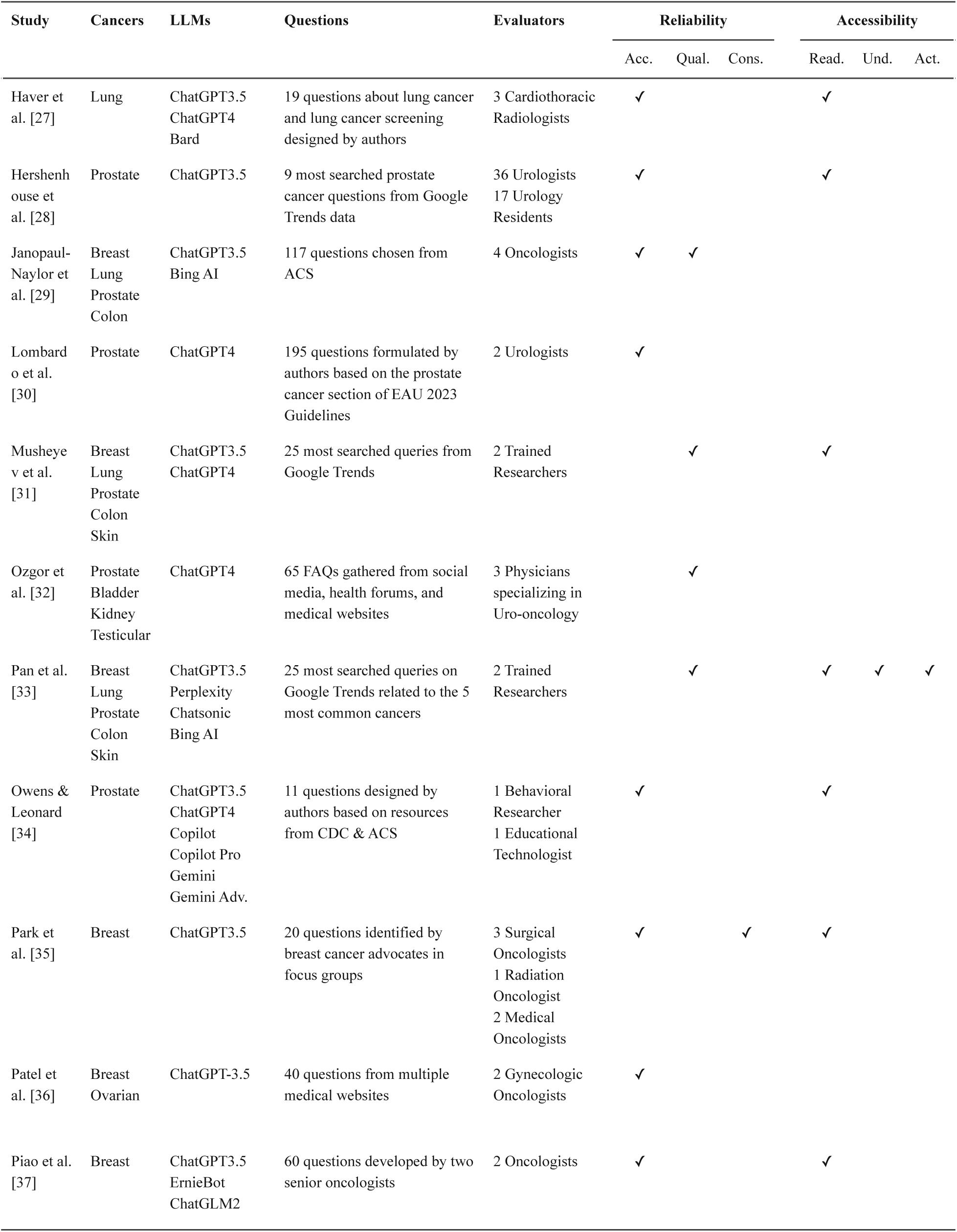
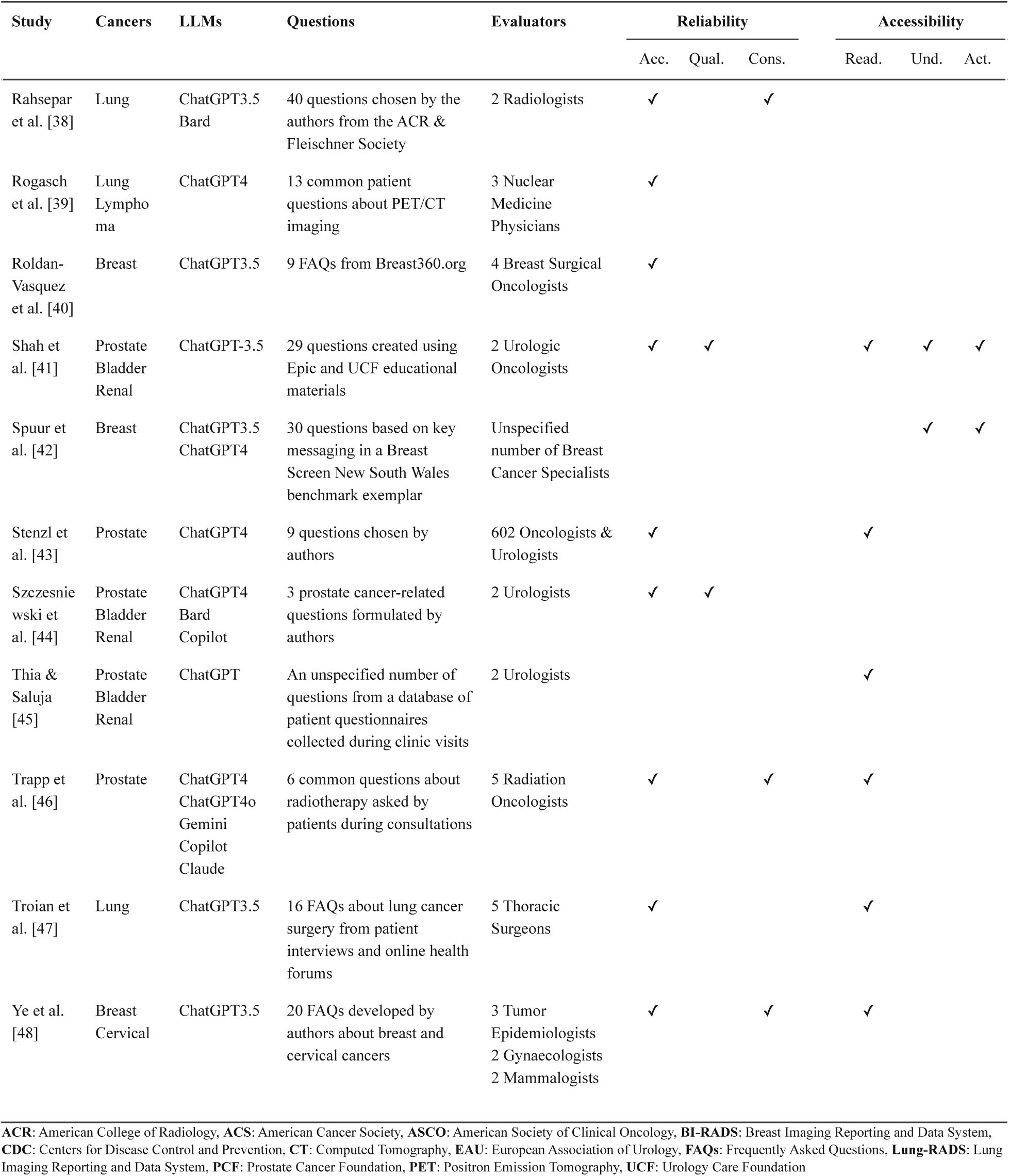
Overview of studies included in the review. Reliability measures included accuracy (Acc.), quality (Qual.), and consistency (Cons.). Accessibility measures included readability (Read.), understandability (Und.), and actionability (Act.).

To illustrate the types of questions asked, a representative sample is provided in Table 2. As can be seen in this exemplar, patient inquiries cover a wide range of topics, from general information on the symptoms and causes of each type of cancer, to various screening and treatment options. The interested reader can find the full list of questions for each study in the Multimedia Appendix section of this article (when made available by the authors).

**Table 2.** Sample patient questions used to query LLMs.

| Study | Cancer Type | Patient Questions |
| --- | --- | --- |
| Collin et al. [19] | Prostate | What are the symptoms of prostate cancer?<br>What are the causes of prostate cancer?<br>How is prostate cancer diagnosed?<br>What are the treatments for prostate cancer?<br>What is the survival rate for prostate cancer?<br>What is a PSA test, and why is it done?<br>What are the risk factors for prostate cancer?<br>Can prostate cancer be prevented?<br>How long does it take for prostate cancer to develop?<br>What is the difference between early-stage and advanced prostate cancer? |
| Roldan-Vasquez et al. [40] | Breast | Do I need to make my reconstruction choices before my initial breast cancer surgery?<br>If I need radiation therapy for my cancer, can I have immediate reconstruction?<br>How do breast surgeons work with plastic surgeons for breast cancer patients?<br>Why do I need a surgical biopsy if my core biopsy did not show cancer?<br>Why shouldn't I just have my lump surgically removed to see if it is cancerous?<br>Does my elderly mother really need surgery to treat her breast cancer?<br>If I have a mastectomy can I save my nipples?<br>Shouldn't I have bilateral mastectomies?<br>What are my reconstruction choices after mastectomy? |
| Rogasch et al. [39] | Lung | How long does a PET scan take?<br>Is a PET scan harmful?<br>How should I prepare for a PET scan?<br>I'm a caregiver to a toddler. Are there any precautionary measures after a PET scan?<br>Can I take a PET scan as a diabetic?<br>Is a PET scan recommended for lung cancer before surgery?<br>Why is a PET scan needed for Hodgkin lymphoma?<br>How accurate is a PET scan for lung cancer?<br>Is a PET scan better than a CT scan for lung cancer?<br>Does negative on PET mean that a lung nodule is benign?<br>Does a hypermetabolic lung nodule on PET mean that I have lung cancer?<br>My PET scan showed a hypermetabolic lung nodule. Should I have it removed?<br>What does "Deauville 5" mean in a PET report? |

In all studies, once questions were formulated, they were posed to one or more LLMs, and the AI-generated responses were evaluated by a panel of experts. Table 1 also contains information on the number and background of the expert evaluators who assessed the LLM responses on multiple criteria. These criteria fall broadly into one of two categories: reliability measures (encompassing accuracy, quality, and consistency), and accessibility measures (encompassing readability, understandability, and actionability). In what follows, we summarize the findings of the reviewed studies along each of these six dimensions.

### 3.1. Reliability Measures

#### 3.1.1. Accuracy

Accuracy refers to the correctness of the LLM responses. For patients to trust this new technology, the answers provided to their questions must be free of errors and misinformation. Accuracy metrics were reported in twenty-seven of the reviewed articles (see Table 1). Despite varied methodologies, researchers commonly used Likert scales to assess expert opinions on response accuracy. For example, Chiarelli et al. [18] used a 3-point Likert scale to label each LLM response as either inaccurate (score = 0), partially accurate (score = 1), or accurate (score = 2). Belge Bilgin et al. [16] used a 4-point Likert scale, where scores from 1 to 4 represented completely incorrect, partially correct, mostly correct, and completely correct answers, respectively. Roldan-Vasquez et al. [40] used a 5-point Likert scale where 1 represented complete inaccuracy and 5 complete accuracy, while Ye et al. [48] employed the most granular Likert scale with 7 points. To allow for direct comparison of results, we used the following linear transformation scheme designed to standardize Likert scores by mapping them onto a common 0-100 range: 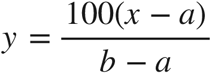.

Here, *x* is the original score, *a* and *b* are the scale’s minimum and maximum values, and *y* denotes the transformed score that lies within the 0-100 range. This approach has been shown to minimize information loss during the transformation of data from its original scale structure [49]. Additionally, the transformed scores can be conveniently interpreted as percentages. In the left panel of Figure 3, we illustrate the distribution of reported accuracy scores post-standardization for n=50 LLMs, noting that some of the twenty-seven studies that measured accuracy evaluated more than one LLM. The normalized accuracy scores ranged from 43.0% to 100%, with a median score of 79.0%.

**Figure 3.**
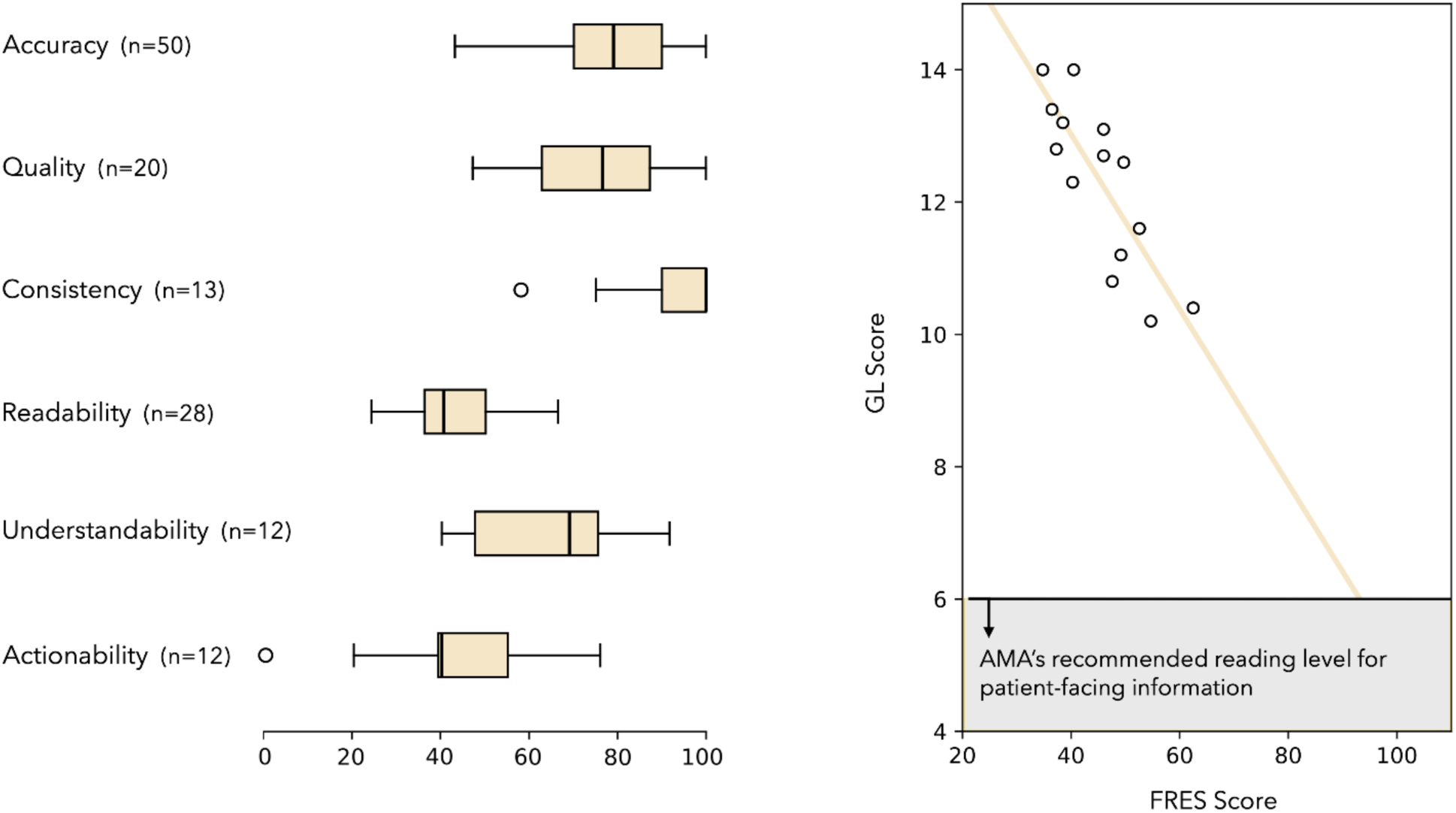
(Left panel) The box plot distribution of accuracy, quality, consistency, readability, understandability, and actionability of LLM responses. (Right panel) The scatter plot of Flesch Reading Ease Scale (FRES) scores and corresponding Grade Level (GL) scores for studies that reported both metrics.

#### 3.1.2. Quality

Accuracy, while essential, does not fully capture the quality of LLM responses. A factually accurate response may be deemed low-quality if it is not entirely relevant or responsive to the question, omits essential information such as risks and benefits associated with screening or treatment, or manifests biases stemming from the training data. Most reviewed studies which reported on the quality of LLM responses utilized the DISCERN tool [50] — a standardized instrument designed to assess the quality of written health information. DISCERN consists of 15 questions, each scored from 1 (low-quality) to 5 (high-quality), assessing relevance, transparency, comprehensiveness, and lack of bias in the answer provided. A 16th question offers an overall quality rating, also using the same 1-to-5 scale.

Besides DISCERN, other studies measured quality using the Global Quality Scale (GQS) with the following scoring structure: 1 = poor quality, not useful for patients; 2 = poor quality, limited usefulness; 3 = partially useful, but missing key information; 4 = good quality, useful for patients; and, 5 = excellent quality, highly useful for patients [15, 20, 22, 32]. Since both DISCERN and GQS utilize 5-point ordinal scales, their results can be mapped to a 0-100 range using the same linear transformation described previously. The distribution of the reported quality scores post-standardization is shown in the left panel of Figure 3. The normalized quality scores ranged from 47.0% to 100%, with a median score of 76.5%.

#### 3.1.3. Consistency

LLMs may not generate the same response when asked the same question multiple times. This is largely due to the stochastic (non-deterministic) nature of text generation in these models. Specifically, most LLMs use a parameter called temperature to control the level of randomness in their output [51]. A higher temperature increases randomness, leading to more diverse responses. In contrast, a lower temperature will result in more deterministic and consistent answers. While a high temperature setting is suitable for creative tasks and applications, it can have a detrimental effect in high-stakes healthcare applications such as answering patient questions.

Six of the reviewed studies evaluated consistency by repeatedly posing the same question to the LLM. Five of these six had experts rate response similarity or consistency [14, 17, 24, 38, 48]. The remaining study [35] used the intraclass correlation coefficient (ICC) of word counts as a proxy for consistency between repeated responses. The box plot of normalized consistency scores is shown in the left panel of Figure 3. Excluding one outlier, the normalized consistency scores of LLMs ranged from 75.0% to 100%, with a median consistency score of 100%.

### 3.2. Accessibility Measures

#### 3.2.1. Readability

Readability refers to how easily LLM responses can be read and understood by patients. Even a reliable model, one which consistently produces accurate and high-quality responses, would be ineffective if the responses were not easily readable. In this context, readability is commonly measured via the Flesch Reading Ease Scale (FRES), which calculates readability based on the average sentence length and the average number of syllables per word [52]. FRES scores range from 0 to 100, with larger scores indicating better readability: 90–100 = very easy to read; 80–89 = easy to read; 70–79 = fairly easy to read; 60–69 = plain English; 50–59 = fairly difficult to read; 30–49 = difficult to read; and, FRES scores below 30 = very difficult to read.

The FRES score distribution of n=28 LLMs in fifteen studies which calculated this readability measure is plotted in the left panel of Figure 3. The reported FRES scores ranged from 24.0% to 66.4%, with a median score of 40.4%. Beyond FRES, researchers also assessed readability using metrics such as the Flesch-Kincaid grade level [53], the Gunning Fog index [54], the automated readability index [55], the Coleman-Liau index [56], and the Simplified Measure of Gobbledygook [57]. These metrics, despite differing computational methodologies, all aim to estimate the U.S. grade level required for understanding written information. For instance, a Flesch-Kincaid grade score of 8 indicates that the text is readable for someone with an eighth-grade education level.

Figure 3 (right panel) displays a scatter plot comparing FRES scores and grade level (GL) scores from studies that reported both. When studies reported more than one GL score, we followed the approach recommended by Haver et al. [27] and averaged them. The American Medical Association (AMA) recommends that consumer health information be written at or below a sixth grade reading level to ensure accessibility for all patients, regardless of their education or literacy level [58]. As can be seen in the figure, all LLMs fell considerably outside this recommended region (highlighted in gray). Unsurprisingly, a fairly strong negative correlation was observed between the FRES and GL scores of LLM responses (R-squared = 0.69).

#### 3.2.2. Understandability

The Patient Education Materials Assessment Tool (PEMAT) is a validated instrument designed to assess the effectiveness of patient-facing health education materials [59]. It offers two distinct versions for printable (PEMAT-P) and audiovisual (PEMAT-A/V) formats. The PEMAT-P section of the instrument includes an understandability component focused on measuring patients’ comprehension of key messages via 19 yes/ no questions, which are scored and reported as a percentage. The left panel of Figure 3 includes the box plot distribution of PEMAT-P understandability scores from six studies (n=12 LLMs) which measured understandability as part of their assessment. The reported understandability scores ranged from 40.0% to 91.7%, with a median score of 69.0%.

#### 3.2.3. Actionability

The second component of the PEMAT-P instrument includes seven yes/no questions designed to determine the ability of patients to identify actionable steps in the provided content. Akin to understandability scores, actionability scores are also calculated as the percentage of positive responses to the seven binary questions. As displayed in the left panel of Figure 3, excluding one outlier, the actionability scores ranged from 20.0% to 76.0%, with a median score of 40.0%.

## 4. Discussion

Our review focused on evaluating the performance of LLMs in addressing patients’ oncology questions, across these six dimensions: accuracy, quality, consistency, readability, understandability, and actionability. As shown in Figure 3, accuracy and quality scores exhibit roughly similar distributions, with median values of 79.0% and 76.5%, respectively. Although these numbers suggest that LLMs generally provide reliable information, inaccurate and low-quality answers do occasionally occur. Such inaccuracies range from minor oversights to significant errors. For example, Coskun et al. [20] highlighted an instance where the LLM responded by stating that “a normal PSA level [is] considered to be 4 ng/mL or lower”. This statement is generally correct but fails to account for individual factors (such as age and prostate volume) that can affect a patient’s normal PSA level. Piao et al. [37] documented another instance wherein ChatGPT incorrectly stated that “surgery can ensure the complete removal of all cancer cells in the patient’s body”.

In contrast to these generally correct but imprecise statements, some LLM answers were entirely incorrect or misleading. For example, Rahsepar et al. [38] and Piao et al. [37] respectively reported LLM responses containing non-existent lung (e.g., Lung-RADS 5) and breast (e.g., BI-RADS 7) imaging categories. In another study, Spuur et al. [42] found several instances of fabricated citations in LLM outputs, likely resulting from the hybridization of real references. This undesired generation of false information by LLMs is termed “hallucination” in computer science literature [60]. To avoid using anthropomorphic language in medical contexts, “fact fabrication” might be a more suitable term to describe this phenomenon [61].

Irrespective of terminology, fact fabrications or hallucinations present a substantial risk, particularly in healthcare applications. Mitigating hallucinations remains an active area of research, with several strategies demonstrating great promise. One such strategy is supervised fine-tuning where a pre-trained LLM’s parameters are fine-tuned using a smaller, task-specific dataset to optimize model performance [62]. Using Retrieval-Augmented Generation (RAG), the accuracy and quality of LLM responses can be improved by incorporating information from external, authoritative sources such as medical textbooks, clinical guidelines, and consensus statements [63]. Chain-of-thought prompting is another strategy that encourages the LLM to explain its reasoning process in a step-by-step sequence, making it easier to identify potential errors in the model’s logic [64].

The third dimension of reliability was consistency, or the ability of LLMs to provide consistent answers when prompted multiple times. Some researchers referred to this concept as stability [14] or reproducibility [17]. With a median consistency score of 100%, our results suggest that LLMs generally perform well in this area (see Figure 3). However, given that only a small fraction of the reviewed studies (6 out of 36) assessed consistency, these results should be interpreted with caution.

In terms of readability, all LLMs exhibited suboptimal performance, as shown in the right panel of Figure 3. Our results indicate that LLM outputs often use complex language which surpasses the average U.S. adult’s eighth grade reading level [65]. Fortunately, there is evidence that prompt engineering, or the deliberate and careful modification of input prompts, can improve the readability of LLM responses. For example, Musheyev et al. [31] reported that adding the phrase “Explain the following at a sixth grade reading level” to the beginning of each prompt lowered the reading level of LLM outputs by two grade levels. Similar substantial improvements in readability have been reported by other researchers through the use of appropriate prompts (see Table 3).

**Table 3.** The effect of prompt engineering on improving the readability of LLM answers.

| Study | Prompt | LLM | Readability<br>(without prompt) | Readability<br>(with prompt) | P-value<br>(less than) |
| --- | --- | --- | --- | --- | --- |
| Haver et al. [26] | Rewrite the following prompt at a 6th grade reading comprehension level ... | ChatGPT-3.5 | FRES=46<br>GL=13.1 | FRES=70<br>GL=8.9 | 0.001<br>0.001 |
| Hershenhouse et al. [28] | Respond to the above patient question in a way that is understandable at or below a 6th-grade level. Be appropriate, accurate, comprehensive, and clear in the response. | ChatGPT-3.5 | FRES=36.5<br>GL=12.8 | FRES=70.2<br>GL=7.4 | 0.0001<br>0.0001 |
| Musheyev et al. [31] | Explain the following at a sixth grade reading level ... | ChatGPT-3.5 | FRES=52.6<br>GL=11.6 | FRES=71.6<br>GL=9.5 | 0.001<br>0.01 |
|  |  | ChatGPT-4 | FRES=62.5<br>GL=10.4 | FRES=75.6<br>GL=8.4 | 0.001<br>0.05 |
| Shah et al. [41] | Explain it to me like I am in sixth grade ... | ChatGPT-3.5 | FRES=38.5<br>GL=12.2 | FRES=66.4<br>GL=7.5 | N/A<br>N/A |
| Owens & Leonard [34] | Explain to someone with low literacy ... | ChatGPT-3.5 | FRES=38.0 | FRES=70.4 | 0.0001 |
|  |  | ChatGPT-4 | FRES=43.1 | FRES=74.1 | 0.0001 |
|  |  | Copilot | FRES=50.8 | FRES=65.1 | 0.0005 |
|  |  | Copilot Pro | FRES=61.2 | FRES=78.8 | 0.0001 |
|  |  | Gemini | FRES=55.7 | FRES=81.0 | 0.0001 |
|  |  | Gemini Adv. | FRES=66.4 | FRES=79.5 | 0.001 |

**FRES:** Flesch Reading Ease Scale, **GL:** Grade Level

With median scores of 69.0% and 40.0%, respectively, understandability and particularly, actionability of LLM responses were lower than desired. It should be noted, however, that the PEMAT-P assessment tool employed to measure understandability and actionability, was not originally designed for evaluating AI-generated content. For instance, PEMAT-P includes several questions about layout, design, and inclusion of visual aids. LLM responses, being purely text-based, may be unfairly penalized on these criteria, contributing to low understandability and actionability scores. Therefore, it is necessary and timely to establish standardized methods and guidelines for evaluating LLMs in medical settings. The Chatbot Assessment Reporting Tool (CHART) is an initiative aimed at filling this gap, specifically for LLMs tasked with providing clinical advice [66].

Our review revealed some deficiencies and limitations that future studies in this area should address. First, the expert evaluation of LLM responses was rarely blinded. In most cases, evaluators knew the responses were AI-generated, and this awareness could have influenced their grading. To mitigate this risk of bias, we recommend that future studies, when feasible, mix human-generated and AI-generated responses in a blinded evaluation. Second, almost all studies used physicians exclusively to evaluate LLM responses. While this is entirely appropriate and necessary for assessing medical accuracy, patients are better suited to judge the accessibility of the answers, as they are the technology’s end-users in this scenario. Future studies should consider recruiting patients with diverse backgrounds and education levels to better assess the patient-centric measures of LLM performance. Patients’ attitudes, perspectives, and feedback could also inform prompt engineering and guide model development. Ideally, randomized controlled trials (RCTs) should be used to test the true efficacy of LLMs, in which real patients pose their questions to chatbots in a controlled environment. In a recent RCT study, Baumgärtner et al. [67] reported a significant reduction in the patients’ information needs following the use of the PROState cancer Conversational Agent (PROSCA). This study was not included in our review because the chatbot PROSCA does not qualify as an LLM, as it only provides fixed and pre-vetted responses. Finally, personalizing LLM interactions by integrating patients’ individual information into the prompt could be a key focus area for future investigations. Without such personalization, LLMs can only offer generalized, one-size-fits-all advice, which may not be suitable for a patient based on their unique medical history or pre-existing conditions.

## 5. Conclusion

Our review of the data suggests that significant improvements are still needed to realize the full potential of LLMs as a source of information for cancer patients. Specifically, future advancements in LLMs must ensure they deliver personalized, hallucination-free information that is easily readable, understandable, and actionable for patients. Though still maturing, LLM-powered chatbots have already influenced how patients access medical information. As with any disruptive technology, LLMs have also been met with a certain degree of skepticism [68]. Decades ago, similar concerns were expressed about the use of computers in medicine [69–71]. Over time, however, the healthcare community adapted and embraced computers and electronic health records. We posit that the adoption of LLMs will follow a similar pattern.

But, to achieve effective integration of LLMs, the physician’s role relative to AI must be clearly defined. It is important to remember that LLMs are not meant to replace doctors, nor can they, as some have feared or warned [72]. Even if AI were to reach a point where it knows every fact about cancer and can convey them accurately, it could never replicate the unique experiences that physicians gain through direct human interactions with patients [73, 74]. Therefore, LLMs should be envisioned as assistant tools for disseminating high-quality information to patients. Physicians can play a critical role as a human-in-the-loop, providing clinical expertise and oversight during training and validation of LLMs. Using techniques such as Reinforcement Learning from human feedback [75], physicians can actively guide LLMs in learning accurately from data, helping to ensure these models uphold the core medical principle of “first, do no harm.”

## Data Availability

All data produced in the present study are available upon reasonable request to the authors.

## Notes

### Competing Interest Statement

The authors have declared no competing interest.

### Funding Statement

This study did not receive any funding.

